# Validation of cardiac image derived input functions for functional PET quantification

**DOI:** 10.1101/2023.09.29.23296343

**Authors:** Murray Bruce Reed, Patricia Anna Handschuh, Clemens Schmidt, Matej Murgaš, David Gomola, Christian Milz, Sebastian Klug, Benjamin Eggerstorfer, Lisa Aichinger, Godber Mathis Godbersen, Lukas Nics, Tatjana Traub-Weidinger, Marcus Hacker, Rupert Lanzenberger, Andreas Hahn

**Affiliations:** Department of Psychiatry and Psychotherapy, Medical University of Vienna, Austria; Comprehensive Center for Clinical Neurosciences and Mental Health (C3NMH), Medical University of Vienna, Austria; Department of Biomedical Imaging and Image-guided Therapy, Division of Nuclear Medicine, Medical University of Vienna, Vienna, Austria

**Keywords:** image-derived input function, arterial input function, functional positron emission tomography (fPET), [^18^F]2-fluoro-2-deoxy-D-glucose ([^18^F]FDG), 6-^18^F-fluoro-l-dopa (6- [^18^F]FDOPA)

## Abstract

Functional PET (fPET) is a novel technique for studying dynamic changes in brain metabolism and neurotransmitter signaling. Accurate measurement of the arterial input function (AIF) is crucial for quantification of fPET but traditionally requires invasive arterial blood sampling. While, image-derived input functions (IDIF) offer a non-invasive alternative, they are afflicted by drawbacks stemming from limited spatial resolution and field of view. Therefore, we conceptualized and validated a scan protocol for brain fPET quantified with cardiac IDIF.

Twenty healthy individuals underwent fPET/MR scans using [^18^F]FDG or 6-[^18^F]FDOPA, with bed motion shuttling between the thorax and brain to capture cardiac IDIF and brain task-induced changes, respectively. Each session included arterial and venous blood sampling for IDIF validation, and participants performed a monetary incentive delay task. IDIFs from fixed-size regions of the left ventricle, ascending and descending aorta, and a composite of all 3 blood pools (3VOI) plus venous blood data (3VOIVB) were compared to the AIF. Quantitative task-specific images from both tracers were compared to assess the performance of each input function.

For both radiotracer cohorts, moderate to high agreement was found between IDIFs and AIF in terms of area under the curve (r = 0.64 – 0.89) and quantified outcome parameters (CMRGlu and Ki(r)=0.84–0.99). The agreement further increased for composite IDIFs 3VOI and 3VOIVB for AUC(r)=0.87–0.93) and outcome parameters (r=0.96–0.99). Both methods showed equivalent quantitative values and high spatial overlap with AIF-derived measurements.

Our proposed protocol enables accurate non-invasive estimation of the input function with full quantification of task-specific changes, addressing the limitations of IDIF for brain imaging by sampling larger blood pools over the thorax. These advancements increase applicability to virtually any PET scanner and to clinical research settings by reducing experimental complexity and increasing patient comfort.

**Graphical Abstract:** 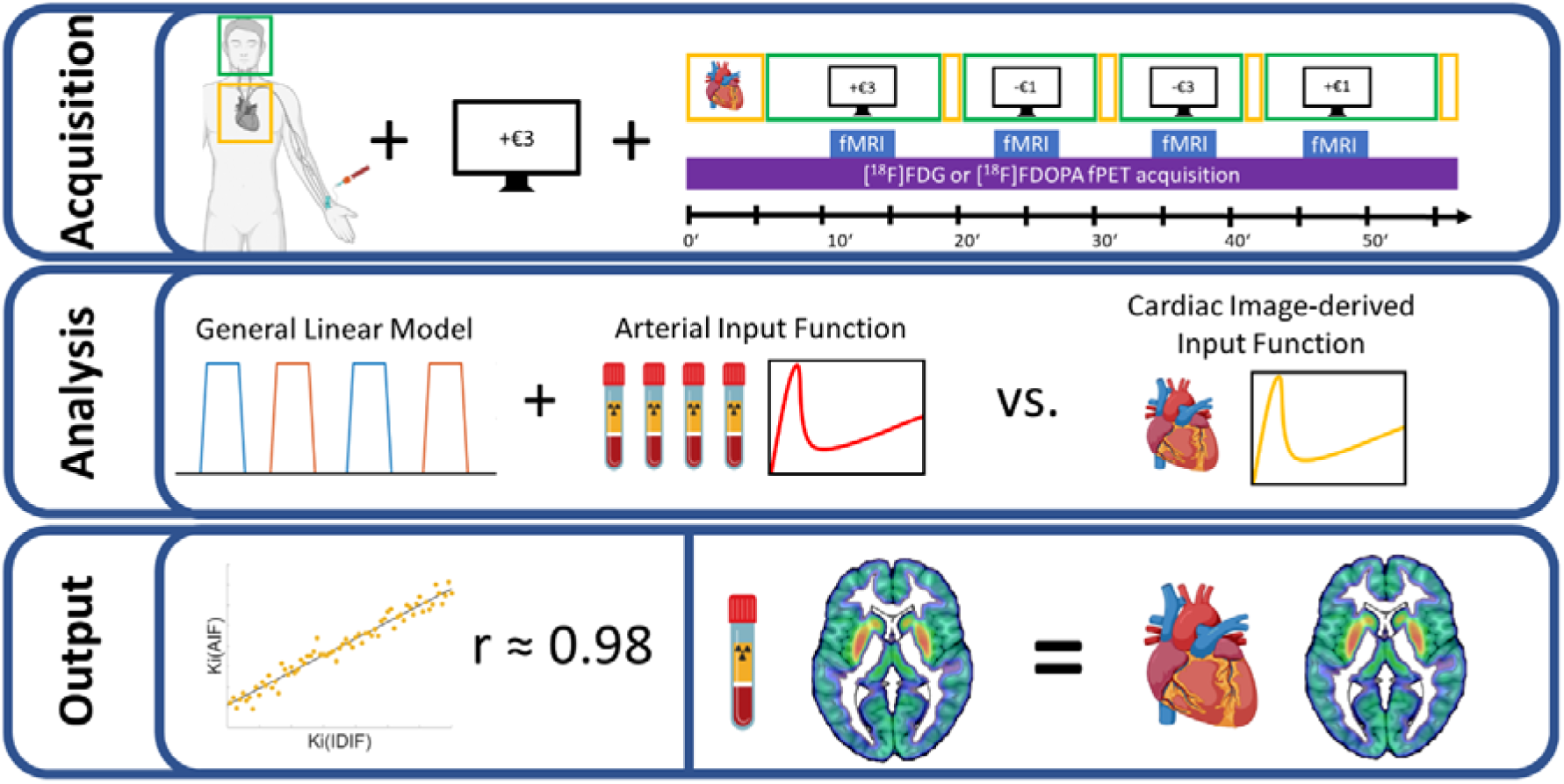

## Introduction

Positron emission tomography (PET) imaging is a widely utilized method for visualizing and quantifying biological processes *in vivo* [1–4]. This includes the quantification of metabolic responses or neurotransmitter signaling during cognitive processing by the recently introduced framework of functional PET (fPET) [5, 6]. Using [^18^F]2-fluoro-2-deoxy-D-glucose ([^18^F]FDG) the approach has successfully identified task-relevant brain networks [3, 7] and revealed decoupling of glucose metabolism and hemodynamic signals [8, 9]. Furthermore, fPET has been utilized to quantify reward-specific changes in dopamine synthesis using 6- ^18^F-fluoro-l-dopa (6-[^18^F]FDOPA) [10].

A pivotal aspect in understanding these physiological and pathological phenomena is the accurate quantification of PET data, which is achieved by dynamic acquisition and the characterization of spatio-temporal patterns of tracer kinetics. Compared to the standard uptake value (SUV), dynamic PET provides more robust outcome parameters [11–13]. However, full kinetic analysis at the voxel level is rarely carried out due to high noise, leading to less reliable parameter estimates and inconsistent models. Simpler graphical modeling methods aim to address these limitation, but in exchange, do not allow the separate estimation of each rate constant [11–13].

The above quantification techniques still rely on measuring the arterial input function (AIF), typically obtained through invasive blood sampling. However, this approach can be challenging and impractical, requires skilled staff and additional medical effort, particularly in subpopulations where arterial access is compromised. A non-invasive alternative is the image-derived input function (IDIF), which extracts the input function from a blood pool within the PET images [14]. In brain studies, the approach has not gained widespread application for several reasons. These include the limited spatial resolution of PET scanners, which lead to spill-over of activity from/to adjacent tissues, potentially affecting the accuracy and reliability, especially for small blood pools within the field of view (FOV) [15]. Particularly the carotid arteries are prone to image noise and artifacts caused by patient motion, scanner instabilities, and low photon counts [15]. In contrast, larger blood pools in the thorax, such as the left ventricle and aorta, offer a more stable and accurate IDIF estimation [14], but these are not within the FOV when imaging the brain with conventional scanners.

This study introduces a novel approach for a minimally invasive fPET scanning protocol to quantify metabolic changes or neurotransmitter synthesis using image-derived input functions (IDIFs). By employing stop-and-go bed motion on a conventional scanner system, the FOV is alternated between the thorax and brain, enabling the acquisition of both the IDIF and the brain response to cognitive processing, respectively. To allow generalizability, the approach is carried out for the quantification of glucose metabolism with [^18^F]FDG and dopamine synthesis with 6-[^18^F]FDOPA. IDIFs extracted from thoracic blood pools are validated with AIFs for both radioligands with respect to input function characteristics and final outcome parameters of net influx constants.

## Materials and Methods

### Participants

Twenty-one healthy individuals were recruited and underwent a single fPET/MRI examination. Participants were injected with either [^18^F]FDG (age: 21±1 years, 3/10 female) or 6-[^18^F]FDOPA (age: 24±4 years 4/10 female). One participant was excluded due to a failure of the automatic blood sampling system. All participants underwent a standard medical examination at the initial screening visit. After detailed explanation of the study protocol, all participants gave written informed consent. Participants were insured and reimbursed for their participation. The study was registered in EudraCT (2019-004880-33) and approved by the Ethics Committee (ethics numbers: 2259/2017 and 2321/2019) of the Medical University of Vienna and procedures were carried out in accordance with the Declaration of Helsinki.

### Cognitive Task

To examine reward and punishment processing, we employed a modified version of the well-established monetary incentive delay (MID) task. Participants were tasked with maximizing reward and minimizing loss by responding to stimuli within specific time limits. The task included 2 win and loss blocks (297s each), with reaction time limits adjusted to manipulate the probability of block outcomes. Prior to the fPET scan, each participant’s individual reaction time was measured. Within each block, the probability of monetary gain and loss was manipulated by adjusting the reaction time limit by±50ms. This resulted in two blocks associated with higher potential monetary gains and two blocks associated with higher potential monetary losses. During baseline phases, subjects were instructed to look at a crosshair, stay awake and let their minds wander. For a comprehensive description of the adapted MID implementation, please refer to the work by Hahn et al.[10].

### PET/MRI data acquisition

Synthesis of both tracers was performed each measurement day. Participants injected with 6-[^18^F]FDOPA received both 150mg Carbidopa and 400mg Entacapone approximately 1 hour prior to tracer application. This was done to block the peripheral metabolism of the radioligand by amino acid decarboxylase and catechol-O-methyl transferase [16, 17]. Both radioligands were administered simultaneously with fPET start using a bolus + constant infusion protocol, as described previously [3, 14] (supplement).

fPET data was collected in list-mode using a stop-and-go bed movement strategy to alternate between brain and thorax regions. The fPET scan started over the thorax, allowing for the determination of image-derived input functions (IDIF) from the left ventricle, ascending, and descending aorta. After 6 min, the bed moved to the brain field of view to acquire baseline and MID task data (4x5min) in a block design. After each task block, the bed returned to the thorax to acquire additional IDIF data points (4x30s). This process was repeated multiple times to obtain reliable IDIF, baseline, and task data (figure 1, supplement).

**Figure 1:**
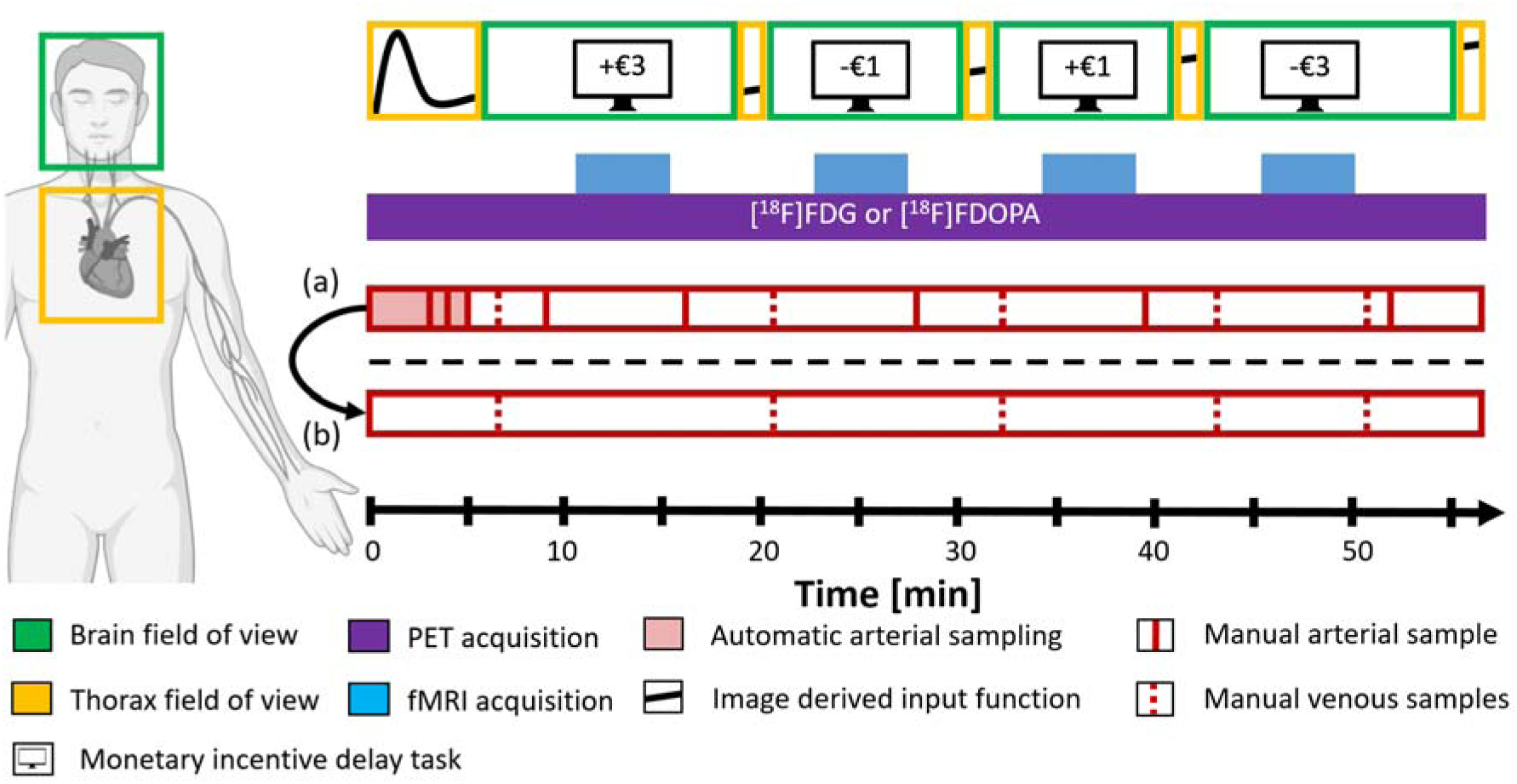
Graphical overview of the measurement protocol. For each participant, the PET scan starts with the PET-FOV placed over the thorax (orange box) for 5:30 min to acquire the initial peak of the tracer. During this time three manual arterial blood samples are taken at minute 3, 4 and 5 (red line). Afterwards, the bed position is moved to the brain (green box) and the start of fPET acquisition is started while the participant views a cross and lets his thoughts wander. At minute 11 the first MID task block begins parallel to the start of fMRI sequence. Before and after task performance, both manual arterial and venous samples are taken. Following the end of the brain block the bed automatically moves to the thorax to acquire further data points for the IDIF in a stop-and-go bed motion. This process is repeated multiple times to provide a robust estimates for both the IDIF and fPET task metrics. (a) depicts the protocol used to validate the IDIFs. (b) indicates the final simplified protocol to calculate fully quantified data.

### Blood sampling and input function construction

Arterial blood samples were drawn from the radial artery and venous samples from the cubital vein (supplement). The AIF was constructed by combining the activities obtained from the automatically and manually collected samples. For IDIFs, fixed-size volumes of interest (VOIs) were manually placed in the left ventricle and thoracic aorta (ascending and descending) using both the mean PET image and structural thorax T1 as reference. For the aorta, a cylindrical VOI with a diameter of 3.3mm and a length of 12mm was used. The left ventricle VOI was defined as a spherical VOI with a diameter of 9.9mm. The mean activity within each VOI was extracted for each time point, representing the IDIFs. In addition, two composite IDIFs were created. These included the left ventricle’s initial time course during the first thorax bed position, which has been shown to be most accurate [14]. Subsequently, the tail of the input function was estimated by linearly fitting a function over all 3 IDIF time courses after 5min, referred to as 3VOI. The second composite IDIF was further included the venous samples to the linear fit, henceforth 3VOIVB.

The resulting input functions were then multiplied with the plasma to whole-blood ratio from the manual blood samples (average for [^18^F]FDG and linear fit for 6-[^18^F]FDOPA). As the intake of carbidopa and entacapone[18] combined with the B+I radioligand administration [10] substantially reduces the amount of radioactive metabolites of 6-[^18^F]FDOPA, a literature-based correction was used (figure S3). The fraction of 6-FDA was fitted with a single exponential function and converted to match the B+I protocol. Additionally, we conducted simulations to investigate the potential biasing effects of increasing the 6-FDA metabolite by 10%, 20%, 30%, and 40% on the quantification process. To ensure alignment with the PET frames, all input functions were linearly interpolated.

### Processing data

Each list-mode PET block was reconstructed using the ordinary Poisson ordered subset expectation maximization algorithm (3 iterations, 21 subsets), see supplement for framing and correction methods. The thorax and brain frames were concatenated separately and decay corrected to the start of the measurement. Brain data was preprocessed using SPM12 (Wellcome Trust Centre for Neuroimaging), as described previously [3] (supplement).

A general linear model was utilized to extract task effects from baseline metabolism. The model included task regressors for win and lose blocks. Additionally, PCA components of the six motion parameters that explained more than 90% of variance were added as motion regressors. The baseline was defined as average across all grey matter voxels, excluding those active during fMRI task performance (contrast success > failure, p<0.001 uncorrected) and those identified in a meta-analysis of the MID task (contrasts reward/loss anticipation and reward outcome) [10]. One frame before and after bed movement was deweighted to 0.5 to reduce potential effects induced by the bed movement. The Gjedde-Patlak plot was used to estimate voxel-wise CMRGlu and net influx constant Ki for [^18^F]FDG and 6-[^18^F]FDOPA respectively. The slope was fitted from t*=20 min after infusion start.

### Statistical analysis

To evaluate the similarity between the IDIFs and the gold standard AIF, we conducted linear regression analysis and computed Pearson correlation coefficients for both the area under the curve and peak values. Furthermore, regional values of CMRGlu and Ki were extracted using the Harvard Oxford subcortical atlas and cortical regions from the Oldham meta-analysis (reward anticipation and loss anticipation, figure S1) [19]. We then assessed the correlation between outcome parameters obtained from IDIFs and AIF (i.e., CMRGlu values from [^18^F]FDG and 6-[^18^F]FDOPA Ki) for both win and loss conditions using Pearson’s correlation coefficient.

To determine the equivalency between the AIF and IDIFs on a regional level, we employed two one-sided t-tests [20]. We also tested for potential bias between venous and arterial blood samples using a paired t-test on mean values for the [^18^F]FDG cohort. For the 6- [^18^F]FDOPA cohort, each venous blood sample was interpolated to match the time of the arterial sample, and individual comparisons were made using a paired t-test. Significance level was p<0.05 for all tests.

## Results

### Comparison of input functions

Figure 2 provides a visual representation of the time course of each input function for both [^18^F]FDG (a) and 6-[^18^F]FDOPA (b) cohorts. The peak values observed in the ascending aorta IDIF were significantly higher than those of all other IDIFs and the AIF (p<0.0001). However, there were no significant differences in peak values between the AIF and the other IDIFs (all p>0.11). The AIF exhibited a later peak (mean±SD: 110±13 s) compared to the ascending aorta (mean±SD: 56±5 s), descending aorta (mean±SD: 72±5 s), left ventricle, 3VOI, and 3VOIVB (mean±SD: 71±6 s).

**Figure 2:**
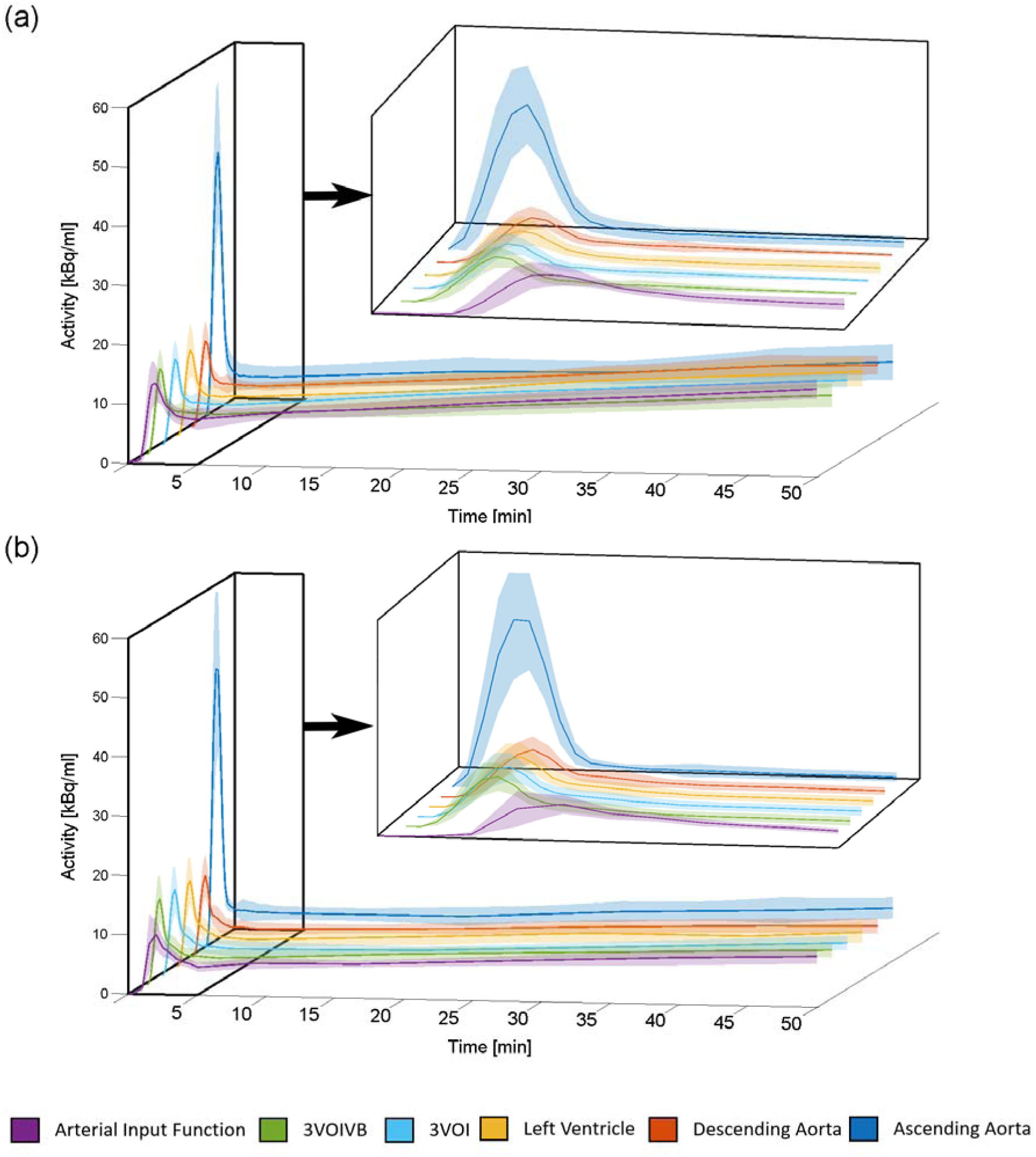
Graphical overview of all input functions. The mean and standard deviation of each input function’s time course for the [^18^F]FDG (a) and 6-[^18^F]FDOPA cohort (b). The top right inlay represents the first 5 minutes of the entire time course. The ascending aorta displays a much higher peak than all other input functions. Moreover, the arterial input function (AIF) peaks later than all image derived input functions. Finally, both the 3VOI and 3VOIVB display the highest similarity to the AIF.

Both cohorts demonstrated comparable degrees of correlations between IDIFs and AIF (table 1). Generally, the left ventricle IDIF showed the highest similarity with the AIF (AUC(r)=0.83-0.89). The match with the AIF increased for the combined IDIF (3VOI(r)=0.87–0.91) and even more when combined with the venous blood samples (3VOIVB/(r)=0.92– 0.93).No significant difference in [18]FDG plasma to whole-blood ratio was found between arterial and venous samples (p=0.82, figure 3a). However, significant underestimation in venous 6-[^18^F]FDOPA concentration was observed at multiple time points (p<0.02). After including an additive factor of 0.0496, no signification differences remained (p>0.69, figure 3b).

**Figure 3:**
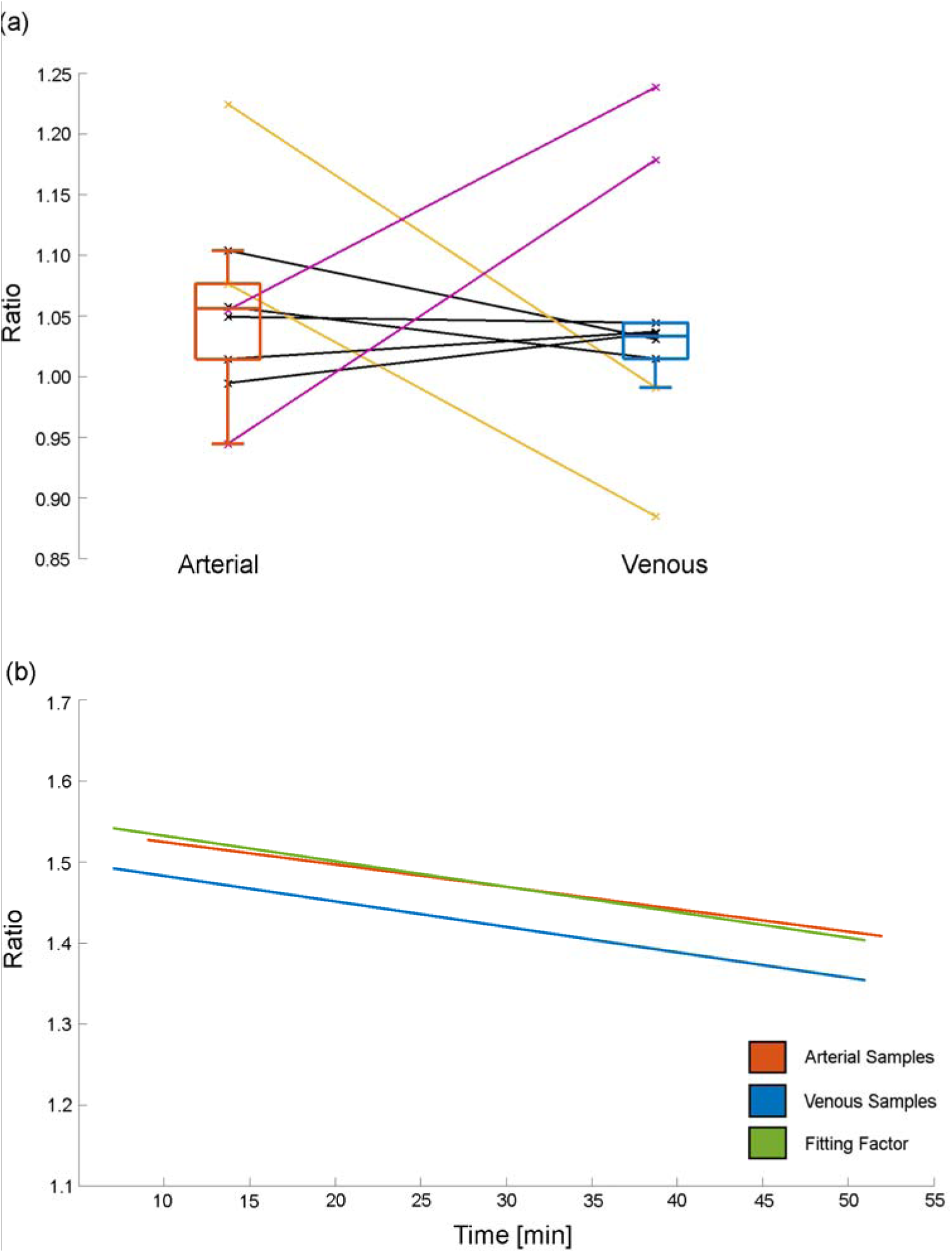
Comparison of arterial and venous plasma to whole-blood ratios for [^18^F]FDG (a) and 6-[^18^F]FDOPA (b). While (a) shows no significant difference between arterial and venous samples, a couple of outliers were present. The yellow outliers’ late venous samples were not collected resulting in an underestimation. In comparison, purple initial venous samples were not collected. All black lines represent participants were all blood samples were taken. (b) In the case of 6-[^18^F]FDOPA, a linear underestimation from the venous samples can be seen (blue) when compared to arterial (orange). By applying a fitting factor (green), estimated over all 10 participants (add=0.0496), this bias can be corrected.

**Table 1:** Comparison of each image derived input function to the gold standard, arterial input function for both [^18^F]FDG and 6-[^18^F]FDOPA tracers. Both the area under the curve and peak values are compared. Bold values indicate the highest correlations per parameter and tracer.

| VOI | [18F]FDG |  | [18F]FDOPA |  |
| --- | --- | --- | --- | --- |
|  | AUC | Peak | AUC | Peak |
| Ascending Aorta | 0.82 | 0.61 | 0.523 | **0.858** |
| Descending Aorta | 0.64 | **0.79** | 0.825 | 0.66 |
| Left Ventricle | 0.83 | 0.60 | 0.892 | 0.816 |
| 3VOIF | 0.91 | 0.60 | 0.866 | 0.816 |
| 3VOIFVB | **0.92** | 0.60 | **0.930** | 0.816 |

### Input function effects on quantified values

The highest correlations of regional CMRGlu and Ki values were found between the AIF and both 3VOI and 3VOIVB IDIFs (r=0.957–0.998, table 2). Accordingly, CMRGlu and Ki values obtained using both the 3VOI and 3VOIVB IDIFs in both cohorts were equivalent to those quantified using the AIF (all p<0.02, table 3). However, regional values derived from the 3 thoracic blood pools were not equivalent to the AIF (p>0.1). Additionally, the magnitude of CMRGlu and Ki values induced by the task in both win and loss conditions were consistent with those reported in previous literature [5, 10].

**Table 2:** Correlation analysis of [^18^F]FDG CMRGlu and 6-[^18^F]FDOPA Ki over all participants between image derived and arterial input functions. *Cortical regions extracted from the Oldham meta-analysis of the monetary incentive delay task [21]. Bold values represent the best correlation for each tracer and region.

| Tracer | Input Function | Brain Regions |  |  |  |
| --- | --- | --- | --- | --- | --- |
|  |  | Accumbens | Caudate | Putamen | Oldham* |
| **[18F]FDG** | Ascending Aorta | 0.955 | 0.903 | 0.938 | 0.989 |
|  | Descending Aorta | 0.984 | 0.983 | 0.984 | 0.989 |
|  | Left Ventricle | 0.988 | 0.980 | 0.987 | 0.992 |
|  | 3VOIF | 0.993 | 0.992 | **0.988** | 0.997 |
|  | 3VOIFVB | **0.995** | **0.992** | 0.987 | **0.998** |
| **[18F]FDOPA** | Ascending Aorta | 0.907 | 0.879 | 0.841 | - |
|  | Descending Aorta | 0.984 | 0.969 | 0.971 | - |
|  | Left Ventricle | 0.983 | 0.973 | 0.968 | - |
|  | 3VOIF | **0.988** | **0.975** | **0.974** | - |
|  | 3VOIFVB | 0.985 | 0.957 | 0.965 | - |

**Table 3:** Regional equivalency tests between each image derived input function and arterial input function (significance indicates equivalence). Cohen’s d represents the standardized mean difference between the AIF and each IDIF. *Cortical regions extracted from the Oldham meta-analysis of the monetary incentive delay task [19].

| Tracer | Input Function | Brain Regions |  |  |  |  |
| --- | --- | --- | --- | --- | --- | --- |
|  |  | Cohen's d | Accumbens | Caudate | Putamen | Oldham* |
| **[18F]FDG** | 3VOIF | 0.33 | <0.0001 | 0.0072 | <0.0001 | 0.0035 |
|  | 3VOIFVB | 0.22 | 0.0001 | <0.0001 | <0.0001 | 0.0266 |
| **[18F]FDOPA** | 3VOIF | 0.69 | 0.0001 | 0.002 | 0.002 | - |
|  | 3VOIFVB | 0.048 | 0.0007 | 0.0296 | 0.0307 | - |

Simulations were conducted to assess the impact of increased 6-FDA metabolite fractions on Ki values. The results showed subtle changes, with a 10% group increase in 6-FDA resulting in a 0.85% increase in task-specific Ki values across ROIs. Subsequent simulations with 20%, 30%, and 40% increases demonstrated corresponding increases of 2.2%, 5.2%, and 8.4%, respectively. According to Ishikawa et al., a variation of up to 30% is plausible [18].

## Discussion

The aim of the study was to compare cardiac IDIFs to the gold standard AIF and assess the validity of derived quantitative values in a fPET framework for multiple tracers. The results demonstrated a variable agreement of the different IDIFs when compared to the AIF. While the IDIFs extracted from a single blood pool showed a moderate to high match in peak values and AUC, the shape of the input function varied at certain time points (figure S2), which influenced the accuracy of task quantification. Both composite IDIFs without (3VOI) and with venous blood samples (3VOIVB) exhibited excellent agreement for input function and outcome parameters, with regional quantified values being equivalent to those derived from the AIF. Moreover, the performance of the IDIFs was statistically equivalent to the AIF for both radiotracers [^18^F]FDG and 6-[^18^F]FDOPA, indicating generalizability of the approach. This allows to derive a simplified protocol, without requirement of arterial blood samples (figure 1b). Taken together, these advancements increase applicability to virtually any PET scanner and also clinical research settings by reducing experimental complexity and increasing patient comfort.

Next-generation PET/CT scanners boast an improved spatial and temporal resolution and a greater field of view which improves the choice of blood pools, the differentiation of tissue types and subsequent IDIF extraction [14, 21]. However, even with these improvements, the IDIF extraction from the typical arterial pools has proven to be difficult. Small vessels such as the carotid, brachial, and femoral arteries are still affected by dispersion and partial-volume effects due to their size [22]. Utilizing larger blood pools in the thorax can help alleviate these problems and improve accuracy [21]. Our results highlight that an accurate IDIF extraction from thoracic blood pools can also be performed on widely available PET/MR or PET/CT scanners. The agreement with the AIF gold standard is even further increased by modelling the time course of multiple blood pools.

While the use of larger thoracic blood pools provides more accurate IDIF estimation, their acquisition with small FOV scanner (e.g., with stop-and-go as well as continuous bed movement) limits quantification to graphical approaches, which only allows for the estimation of the net influx constant. These graphical methods do not require that the shape of the initial part of the input function be precisely estimated, as they mainly rely on the area under the curve. Thus, graphical methods are less affected by IDIF errors and in this context provide more robust quantification than compartmental modeling [15]. Despite their potential, graphical methods have been shown to be potentially susceptible to bias [23], primarily influenced by the accuracy of estimating the later segments of the input functions. Incorporating blood samples for scaling purposes can greatly improve the accuracy of the IDIF curves’ tail [23]. Our results also suggest that the use of venous samples improves both IDIF shape and quantification accuracy but to a lesser degree than previously reported, indicating that the benefit is dependent on the radiotracer.

The strong performance of the IDIFs for both [^18^F]FDG and 6-[^18^F]FDOPA suggests that the proposed approach is generalizable, at least for radioligands with (partly) irreversible kinetics and subsequent quantification with the Patlak plot. Still, we propose that this may also be successfully extended to quantification of certain reversible radioligands with the Logan plot. Although this graphical approach requires the integral of both blood and tissue activity[24], radioligands with slow tissue kinetics may not require full sampling of the initial part of the time activity curve. On the other hand, most reversible radioligands require determination of radioactive metabolites, usually from blood samples. Here, it is important to acknowledge that arterial tracer kinetics may differ from venous ones [25, 26]. Substitution can only be done if the venous samples are obtained during a period of transient equilibrium. However, the time required to achieve this equilibrium varies for each tracer. This also applies to irreversible radioligands with respect to plasma to whole-blood ratio, e.g., arteriovenous equilibrium for [^18^F]FDG is reached approximately 10 to 15 minutes after injection [27]. This can also be seen in our [^18^F]FDG cohort data, where we found no significant differences between venous and arterial samples. While our 6-[^18^F]FDOPA blood data also seemed to reach arteriovenous equilibrium around the same time window as [^18^F]FDG, there was a constant underestimation in the venous samples. This however, can be corrected for by implementing an additive factor for quantification. Of note, underestimation of venous samples had no significant effect on the final outcome parameters. Here, the advent of long-axial FOV PET/CT scanners, that allow simultaneous recording of the brain response, thoracic IDIF and further organs involved in metabolism, holds promise for a non-invasive full compartmental modeling approach, where rate constants can be estimated without blood sampling. While the peripheral metabolism of 6-[^18^F]FDOPA was blocked by applying Carbidopa and Entacapone, we cannot rule out effects of other metabolites like 6-FDA. As we did not measure metabolite fractions, we were limited to a literature-based correction rather than an individualized approach. While this adjustment might alter dopamine synthesis values to some degree, the metabolism is further reduced by using a bolus + infusion protocol (figure S3) and simulations indicate negligible changes.

In sum, we propose a robust and accurate protocol to quantify task-induced [^18^F]FDG metabolic changes and 6-[^18^F]FDOPA dopamine synthesis without the burden of arterial sampling. This protocol utilizes IDIFs extracted from thoracic blood pools, which was validated with the gold standard AIF. Combining IDIFs across several blood pools showed an excellent match to the AIF. Furthermore, the quantitative images derived from the IDIFs were equivalent to those derived from AIF. Overall, this protocol provides a promising approach to reduce patient burden and experimental complexity while accurately quantifying acute task-specific changes. The approach can be implemented on any PET scanner and offers potential extensions to numerous additional applications

## Acknowledgments

This research was funded in whole, or in part, by the Austrian Science Fund (FWF) [KLI 1006, PI: R. Lanzenberger] and the WWTF Vienna Science and Technology Fund [CS18- 039 Co-PI: Rupert Lanzenberger]. For the purpose of open access, the author has applied a CC BY public copyright license to any Author Accepted Manuscript version arising from this submission. We thank the graduated team members and the diploma students of the Neuroimaging Lab (NIL, headed by R. Lanzenberger) as well as the clinical colleagues from the Department of Psychiatry and Psychotherapy of the Medical University of Vienna for clinical and/or administrative support. The scientific project was performed with the support of the Medical Imaging Cluster of the Medical University of Vienna. Parts of figures were created with BioRender.com.

## Disclosure / Conflict of Interest

With relevance to this work, there is no conflict of interest to declare. R. Lanzenberger received travel grants and/or conference speaker honoraria within the last three years from Bruker BioSpin MR, Heel, and support from Siemens Healthcare regarding clinical research using PET/MR. He is a shareholder of the start-up company BM Health GmbH since 2019. M. Hacker received consulting fees and/or honoraria from Bayer Healthcare BMS, Eli Lilly, EZAG, GE Healthcare, Ipsen, ITM, Janssen, Roche, and Siemens Healthineers.

## Data Availability Statement

Raw data will not be publicly available due to reasons of data protection. Processed data and custom code can be obtained from the corresponding author with a data-sharing agreement, approved by the departments of legal affairs and data clearing of the Medical University of Vienna.

## Notes

### Author Declarations

The study was approved by the Ethics Committee (ethics numbers: 2259/2017 and 2321/2019) of the Medical University of Vienna and procedures were carried out in accordance with the Declaration of Helsinki.

