## Supplementary Material for "Validation of cardiac image derived input functions for functional PET quantification"

***For submission to:***

***medRxiv***

### Supplementary Material

### Correspondence to:
Prof. Rupert Lanzenberger, MD, PD

ORCID: https://orcid.org/0000-0003-4641-9539
Medical University of Vienna, Department of Psychiatry and Psychotherapy, Austria

### Materials and Methods

#### Participants

All participants underwent a standard medical examination at the initial screening visit, which included blood tests, electrocardiography, neurological testing and the Structural Clinical Interview for DSM-IV performed by an experienced psychiatrist. Female participants also underwent a urine pregnancy test at the screening visit and before the PET/MRI scan. Exclusion criteria were current and previous (12 months) somatic, neurological or psychiatric disorders, current and previous substance abuse or psychotropic medication, current pregnancy or breast feeding and previous study-related radiation exposure in the past 10 years.

#### PET/MRI data acquisition

Both radioligands were administered simultaneously with fPET start using a bolus (510 kBq/kg/frame, 1min) + constant infusion (40 kBq/kg/frame, 56min) protocol, with a perfusion pump (Syramed µSP6000 with UniQUE MRI-shield, both Arcomed, Regensdorf, Switzerland) as described previously[3, 14].

Structural MRI of the brain was acquired before fPET using a T1-weighted MPRAGE sequence (TE/TR=4.21/2200 ms, TI=900 ms, flip angle=9°, matrix size=240×256, 160 slices, voxel size=1mm isotropic, TA=7:41 min), which was used for spatial normalization. Whereas, the thorax was acquired using a T1-weigthed STARVIBE sequence (TE/TR=1.44/3050 ms, flip angle=5°, matrix size=320x320, 208 slices, voxel size=1.19x1.19x1.2 mm, TA=5:33 min) and used for IDIF localization.

MID functional data was acquired using an EPI sequence (TE/TR=30/2000 ms, flip angle=90°, matrix size=80×80, 34 slices, voxel size=2.5x2.5x2.5 mm + 0.825 mm gap).

#### Blood sampling

For the first 5.5 minutes, blood was automatically sampled from the radial artery (Twilite II system; Swisstrace) and additional manual samples were obtained at 3, 4 and 5 minutes after tracer administration. Thereafter, during baseline phases, both manual arterial (9, 16.5, 28, 39.5 and 52 min) and venous blood (7, 20, 31.5, 43, 51 min) samples were drawn. After each measurement, plasma was separated from whole blood, and the activity of both was measured using a γ-counter (Wizward2, 3”, Perkin Elmer), which was cross-calibrated to the PET/MR scanner.

#### Processing data

The first thorax block was binned (20x5s, 8x10s, 5x30s) to accurately map the initial tracer kinetics. The remaining brain and thorax blocks were binned into 30s frames. Attenuation and scatter correction were performed for brain blocks using a pseudo-CT approach based on the structural T1, while the thorax blocks were corrected using the DIXON MRAC with a CAIPIRINHA sampling pattern[1]. The distance between the head and thorax field of view was set to 335mm for all participants.

The thorax and brain frames were concatenated separately and decay corrected to the start of the measurement. Brain data was preprocessed using SPM12 (Wellcome Trust Centre for Neuroimaging), as described previously[2]. Briefly, fPET data were corrected for head motion (quality=best, registered to mean) and coregistered to the structural T1. The structural MRI was spatially normalized to MNI space, and the transformation matrix was applied to the coregistered fPET images. Images were smoothed with an 8mm Gaussian kernel, masked to include only gray matter voxels and a low-pass filter was applied with the cutoff frequency set to 2.5min.

fPET data was preprocessed using SPM12 (Wellcome Trust Centre for Neuroimaging) and were corrected for head motion (quality=best, registered to mean) and coregistered to the structural T1. The structural MRI was spatially normalized to MNI space, and the transformation matrix was applied to the coregistered fPET images. Images were smoothed with an 8mm Gaussian kernel, masked to include only gray matter voxels and a low-pass filter was applied with the cutoff frequency set to 2.5min.

#### Supplementary Figures

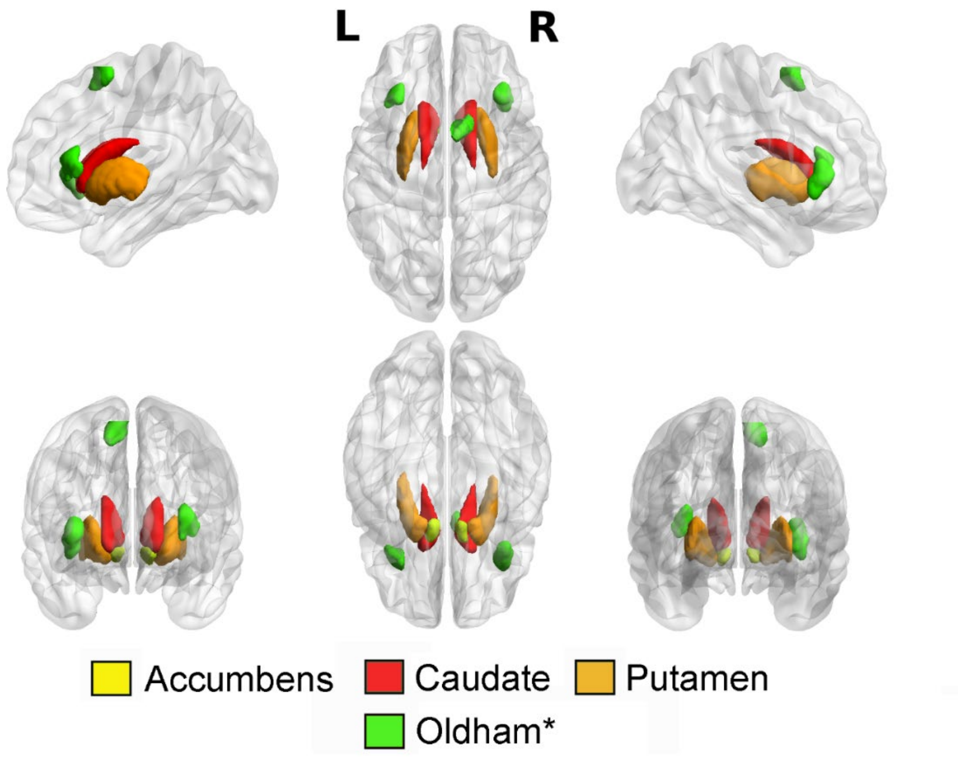


Figure S1: Graphical depiction of the region of interest used in both [$^{18}$F]FDG and 6-[$^{18}$F]FDOPA cohorts. The green (Oldham) cortical regions were extracted from the conjunction analysis of reward and loss anticipation from Oldham et al. [3]. The Oldham regions of interest were not used for 6-[$^{18}$F]FDOPA cohort. Created using BrainNet Viewer [4].

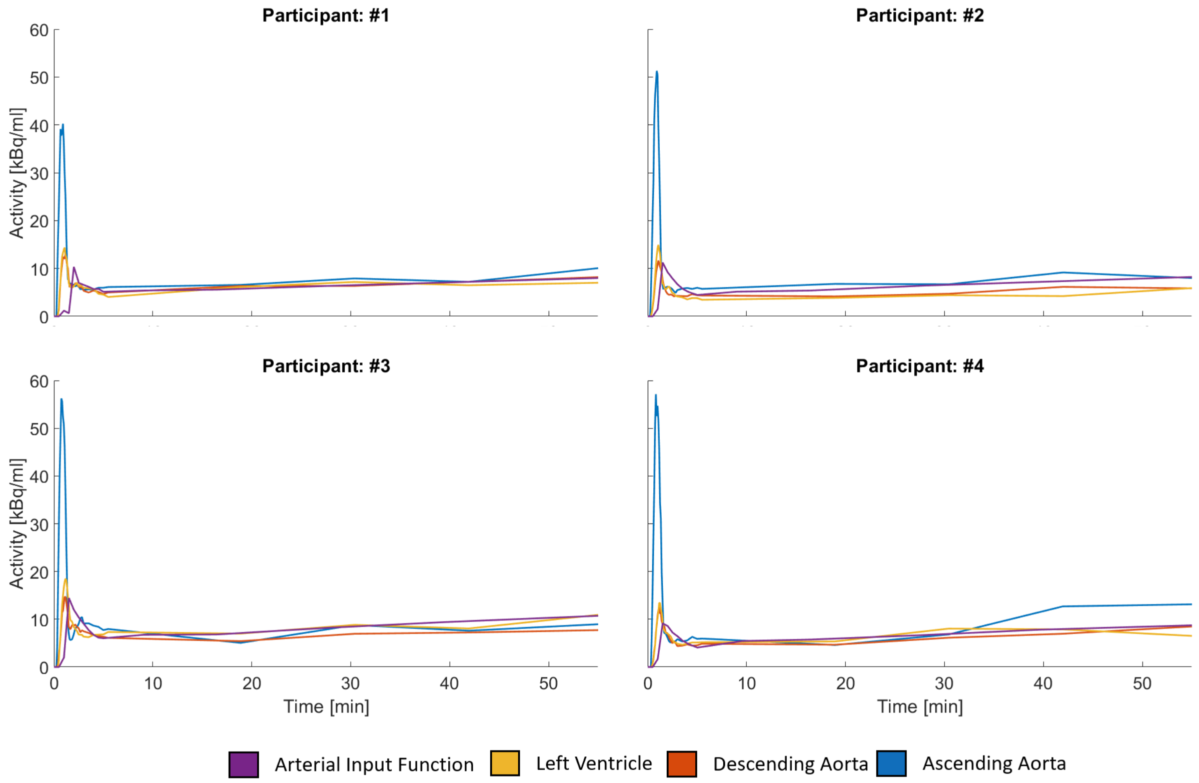


Figure S2: Individual input function time courses of four participants. The shape of the image derived input functions extracted from the left ventricle, ascending and descending aorta varied from the arterial input function. This is more visible during the later stages of the measurement.

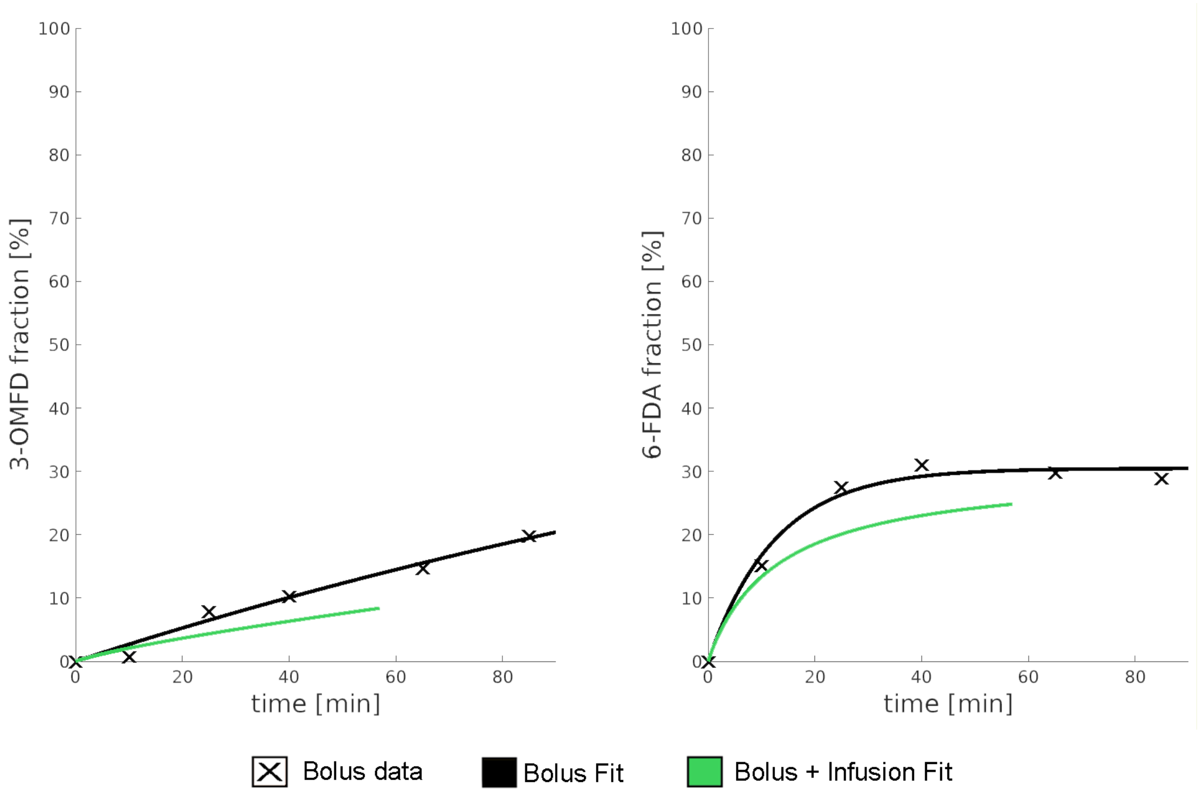


Figure S3: The metabolite fractions (3-OMFD) and (6-FDA) were extracted from previous studies [5, 6] and [7] respectively (black crosses). These data points were then fitted with a single exponential function (black line) and adapted to match the bolus + infusion protocol (green line), showing the reduced metabolism with this protocol. The estimated bolus + infusion 6-FDA fraction data was combined with the individually measured plasma to whole blood ratio to obtain the corrected arterial and image derived input functions. No corrections for the 3-OMFD fraction were done as participants received 150mg Carbidopa and 400mg Entacapone approximately 1 hour prior to tracer application to block the peripheral metabolism of 6-[$^{18}$F]FDOPA.
